# What Additional Insights Does a Bayesian Network Meta-Analysis Provide? Revisiting P2Y12 Inhibitors After Acute Coronary Syndromes

**DOI:** 10.64898/2026.08.31.26361847

**Authors:** Stephen A. Kutcher, Nandini Dendukuri, Sonny Dandona, James M. Brophy

**Affiliations:** Children’s Hospital Eastern Ontario; McGill University Health Centre; McGill UniversityHealth Centre

## Abstract

**Background:** The optimal P2Y12 inhibitor after percutaneous coronary intervention (PCI) remains debated. A recent frequentist network meta-analysis (NMA) concluded prasugrel provided optimal efficacy–safety balance. We update our previous Bayesian NMA with recent trials and contrast our findings with the frequentist NMA.

**Methods:** We extended our 2023 systematic review by adding two trials published after our search cutoff. The primary efficacy endpoint was a composite of all-cause mortality, a recurrent non-fatal myocardial infarction, or non-fatal stroke (MACE). The primary safety endpoint was study-reported major bleeding events. Bayesian network meta-analysis with a primary binomial complementary log-log model with log(time) offset and random effects was performed. A statistical workflow with prior and posterior predictive checks, convergence diagnostics, model comparisons, sensitivity analyses and probabilities for a range of practical equivalence (ROPE: HR 0.90–1.11) are also reported.

**Results:** 19 RCTs (n ≈ 60,619) were identified. For MACE, prasugrel’s probability of a clinical efficacy benefit (hazard ratio (HR) <0.9) was 70% compared to clopidogrel (HR 0.87, 95% credible interval (CrI)0.76–1.03) and 54% compared to ticagrelor. (HR 0.89, 95% CrI 0.73–1.12). For the ticagrelor versus clopidogrel comparison 76% of the posterior probability (HR 0.98, 95%CrI 0.82–1.18) lies in the ROPE. Ticagrelor (HR 1.26, 95%CrI 1.01–1.63) and prasugrel (HR 1.1, 95%CrI 0.88–1.26) showed 86% and 51% probabilities respectively of meaningful bleeding harm (HR > 1.1) versus clopidogrel.

**Conclusions:** Despite 19 RCTs and approximately 60,000 patients, the Bayesian framework revealed clinically important uncertainties and identified probable regions of equivalence among the different P2Y12 inhibitors that were under appreciated with the previous frequentist publication.

## 1. Introduction

Network meta-analysis increasingly informs clinical guidelines, yet different statistical frameworks may lead to different interpretations of the same evidence. We used the contemporary debate regarding P2Y12 inhibitor therapy after acute coronary syndrome to compare the insights provided by Bayesian and frequentist network meta-analysis.

Dual antiplatelet therapy (DAPT) with aspirin and a P2Y12 inhibitor is the cornerstone of secondary prevention after percutaneous coronary intervention (PCI) for acute coronary syndrome (ACS). Three oral P2Y12 inhibitors are available: clopidogrel, ticagrelor, and prasugrel. Current guidelines favour ticagrelor or prasugrel over clopidogrel (1,2), but the evidence base contains important uncertainties that frequentist analyses have not fully captured.

A recent frequentist network meta-analysis(NMA)(3) (15 randomized clinical trials (RCT), n = 48,904) concluded that “prasugrel provided the optimal balance for efficacy and safety”. We revisit the superiority of the available DAPTs by updating our 2023 Bayesian NMA (4) (17 trials, n = 57,814) with two recent trials: TC4 (5), the first dedicated North American RCT comparing ticagrelor and clopidogrel (n = 1,005), and TUXEDO-2 (6), a head-to-head comparison of prasugrel and ticagrelor in diabetic patients with multivessel disease (n = 1,800). The Bayesian approach offers several advantages in this context: it quantifies uncertainty continuously rather than dichotomizing it into significant/non-significant, incorporates prior evidence transparently with explicit sensitivity analysis, distinguishes statistical from clinical significance through the region of practical equivalence (ROPE), and correctly standardizes trials with varying follow-up durations through a complementary log-log model with log(time) offset.

## 2. Methods

Our initial Bayesian NMA followed the PRISMA extension for network meta-analyses(7). Two new RCTs were identified and added to those previously identified in our systematic review (4). The primary analysis included low risk-of-bias RCTs comparing at least two of clopidogrel (C), ticagrelor (T), or prasugrel (P) with a minimum 6-month follow-up, reporting on the hard MACE outcome (all-cause death + non-fatal MI + non-fatal stroke), and at least one reported major bleeding endpoint. Selected RCTs were compared with those identified in the recent frequentist network meta-analysis (3).

### 2.1 Outcomes

The primary efficacy outcome is major adverse cardiovascular events (MACE) a combination of all-cause mortality, non-fatal myocardial infarction, and non-fatal stroke. This “hard” composite was adopted to minimize heterogeneity in outcome definitions across trials, following our previous systematic review and network meta-analysis (4). It differs from the softer primary composites used by some individual trials. For example, in TUXEDO-2 (6), the hard MACE count defined here was a secondary and not a primary endpoint. The safety outcome is trial-reported major bleeding using each trial’s own definition and this heterogeneity in bleeding definitions is noted as a limitation.

All event counts are integers reported at the arm level. An integer continuity correction (0 events → 1 event, n → n+1) is applied only to the two zero-event MACE arms (8,9), giving near-zero rates of 1/(n+1). This is required because the binomial family demands integer responses; the affected trials contribute negligibly to efficacy estimates.

### 2.2 Statistical Model

For each study *i*, arm *j* with *n*_*ij*_ patients and follow-up *t*_*i*_ years, the number of observed utcomes *n*_*ij*_ was modelled as a binomial outcome:

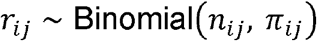

where the event probability *π*_*ij*_ is linked to the linear predictor via the complementary log-log (cloglog) function:

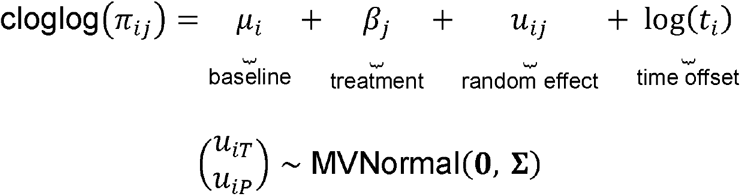

The four components are:

- *μ*_*i*_: the study-specific baseline log-hazard for the reference treatment (clopidogrel) in study *i*. For trials with a clopidogrel arm this is directly informed by observed events. For prasugrel versus ticagrelor trials, it is inferred entirely from the network as these trials contain no clopidogrel arm.
- *β*_*j*_: the population-average log-hazard ratio for treatment *j* (ticagrelor or prasugrel) versus clopidogrel, averaged across the distribution of studies. This is the primary quantity of clinical interest; exponentiating *β*_*j*_ yields the pooled hazard ratio.
- *u*_*ij*_: the study-specific deviation from the population-average treatment effect *β*_*j*_, capturing genuine between-study heterogeneity in treatment response. It is not the mean treatment effect — that role belongs to *β*_*j*_. The bivariate normal rather than independent structure for (*u*_*iT*_,*u*_*iP*_)allows for correlation between study-specific deviations across treatments: a trial population with unusually high thrombotic risk may show greater benefit from both active agents simultaneously. Rather than assuming the two treatment deviations are independent — which would force their correlation to zero by assumption — we place a prior on the correlation matrix **Σ** that is uniform over all mathematically valid correlations, from − 1 to +1 . This lets the data determine whether study-specific deviations for ticagrelor and prasugrel tend to move together or in opposite directions, without imposing a prior belief either way.
- Log(*t*_*i*_): a time offset that converts each trial’s observed cumulative event probability into an annualised hazard rate, placing all trials on a common per-year scale before any parameters are estimated. Without this correction, a trial with 30-month follow-up (TRILOGY ACS, ∼18% cumulative event rate) and a trial with 6-month follow-up (Gasecka, ∼0%) would be pooled as if they measured the same underlying quantity. Its coefficient is fixed at exactly 1 rather than estimated, because this is a mathematical identity arising from the proportional hazards derivation (log(*λt*) = log(*λ*)+ log(*t*)) rather than a free parameter: fixing it at 1 is what makes the linear predictor represent a log-hazard rate. In practice this means the model estimates one common annual hazard rate for each treatment, then each trial’s observed probability is obtained by applying that rate over its own follow-up time.

The cloglog link is the natural choice because it arises directly from the proportional hazards model:

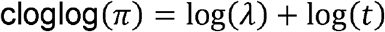

where *λ* is the underlying hazard rate. Exponentiating differences in the linear predictor between two treatments yields hazard ratios (not odds ratios), making the cloglog model preferable to logistic regression when event rates exceed ∼10% or follow-up durations vary — both conditions existing in this network.

### 2.3 Priors

Rather than viewing priors as a weakness of Bayesian analyses, they in fact offer an advantage: they force explicit specification of background assumptions and allow systematic sensitivity testing, in contrast to frequentist analyses where equivalent assumptions are implicit and rarely examined. Our baseline priors were as follows:

**Intercept** *μ*_*i*_ ∼ *normal(-2*.*0, 1*.*0):* baseline log-hazard for the clopidogrel arm (Equation 2). On the event rate scale this covers 1–30%/year, centred on ∼12.7%, consistent with observed control arm rates across trials.

**Treatment effects** *β*_*j*_ ∼ *normal(0, 0*.*5):* log-hazard ratios versus clopidogrel (Equation 2). This weakly informative centred at zero places 95% prior mass on HR [0.37, 2.70], essentially encoding no prior preference for any specific drug.

**Between-study standard deviation (SD)** *τ ∼* *normal(0, 0*.*5)*: internally the model constrains SD parameters to be positive, making this a half-Normal(0, 0.5) distribution. *τ* governs how much the true treatment effect varies from trial to trial — it is the standard deviation of the study-specific deviations *u*_*iT*_ and *u*_*iP*_ in Equation 3. A value of *τ* = 0 would mean all trials estimate exactly the same underlying hazard ratio (no heterogeneity while larger *τ* means the true effect genuinely differs across populations. The covariance matrix **Σ** in Equation 3 is decomposed into these standard deviations (*τ*) and the correlations between them, which are estimated separately from the data. The half-Normal(0, 0.5) prior for *τ* has a prior mean of 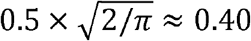 on the log-HR scale.^1^ This implies that study-specific true treatment effects are expected to vary by roughly ± 0.78 log-HR units around the population mean (i.e., ± 1.96 *τ*), corresponding to a 95% prediction interval for any new trial spanning HR [0.46, 2.18]. While wide, this reflects genuine uncertainty about heterogeneity before data are observed. Empirical data on pharmacological cardiovascular interventions suggest typical *τ* values of 0.1– 0.3 on the log-OR scale(10), placing our prior mean of 0.40 on the conservative (wider) end — appropriately cautious for a network spanning diverse populations and geographies.

#### ROPE thresholds

We define HR < 0.90 = clinically meaningful benefit; HR > 1.11 = clinically meaningful harm. While these thresholds are somewhat arbitrary, most clinicians would regard differences within this range as clinically equivalent, and they can be readily adjusted in the post-processing phase. As a supporting justification, almost all cardiovascular RCTs are powered to detect differences that exceed this range, suggesting it represents an acceptable lower bound for clinical relevance. These thresholds match those used in our previous work (4,5).

The influence of these priors was assessed through multiple pre-specified sensitivity analyses (Supplemental Table 1).

### 2.4 Statistical Workflow

The analysis follows a formal Bayesian workflow(11) to verify modelling assumptions and assess robustness. Specifically, we performed (1) prior predictive checks to confirm they imply plausible event rates (2) data simulation with parameter recovery to verify the inference machinery correctly recovers known true parameters; (3) model fitting with convergence diagnostics; (4) posterior predictive checks to assess calibration of fitted models; (5) leave-one-out cross-validation (LOO-CV) for model comparison between random and fixed effects specifications; and (6) the multiple sensitivity analyses. This approach is consistent with the Bayesian analysis reporting guidelines (12), endorsed by the EQUATOR network.

Operationally, models were analysed within the R (13) statistical programming environment using the brms package (14), a high-level front-end interface for Stan (15), a probabilistic programming language that employs Hamiltonian Monte Carlo (HMC) sampling with the No-U-Turn Sampler (NUTS) algorithm for efficient exploration of the posterior distributions. Posterior estimation was performed via the cmdstanr package (16), which interfaces directly with Stan and provides enhanced computational and diagnostic capabilities. Four chains were run with 2,000 warm-up and 4,000 sampling iterations per chain, using adapt_delta = 0.95 and max_treedepth = 12. Convergence was evaluated using the Gelman–Rubin statistic (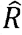, target < 1.01) and bulk effective sample size (target > 400). Posterior summaries are reported as medians with 95% credible intervals (CrI).

The statistical code is available online (https://github.com/brophyj/Bayesian-NMA).

## 3. Results

### 3.1 Trial Selection and Reconciliation

Two new trials were published since our previous systematic review(4) and are included in this analysis.

#### TC4 (5)

Single-centre North American, pragmatic cluster-RCT of ticagrelor versus clopidogrel in ACS patients undergoing PCI. This is the only dedicated North American RCT evidence on this comparison since PLATO(17). The TC4 trial used the 2023 BNMA(4) as its “summary prior,” completing a Bayesian updating cycle.

#### TUXEDO-2 (6)

Multicenter trial from India with a highly selected population of 100% diabetic patients undergoing multivessel PCI (85%), with a subpopulation of chronic coronary syndrome patients (21.8%), comparing prasugrel versus ticagrelor.

Three trials included in this analysis were absent from the JAMA NMA(3): TC4(5) (ticagrelor versus clopidogrel, n=1,005), TREAT(18) (ticagrelor versus clopidogrel, n=3,799), and REDUCE-MVI(19) (prasugrel versus ticagrelor, n=110). TREAT(18) was excluded as it was a post-fibrinolysis, non-PCI population; no reasons were provided for the other exclusions. Conversely, three trials in the JAMA NMA(3) were excluded from our analysis, two for risk of bias concerns(20,21) and one for its one month follow-up(22) falling below our 6-month minimum.

Our main MACE analysis included 19 trials with 60,619 randomized patients (5,6,8,9,17–19,23–34) and the treatment network is displayed in Figure 1. Key inclusions, exclusions and comparisons with the previously published network meta-analyses(3,4) are summarized in Table 1. Individual trial details concerning treatment arms, follow-up duration, reference and comparator raw MACE events are shown in Table 2. The corresponding individual trial major bleeding data is shown in Table 3.

**Table 1:**
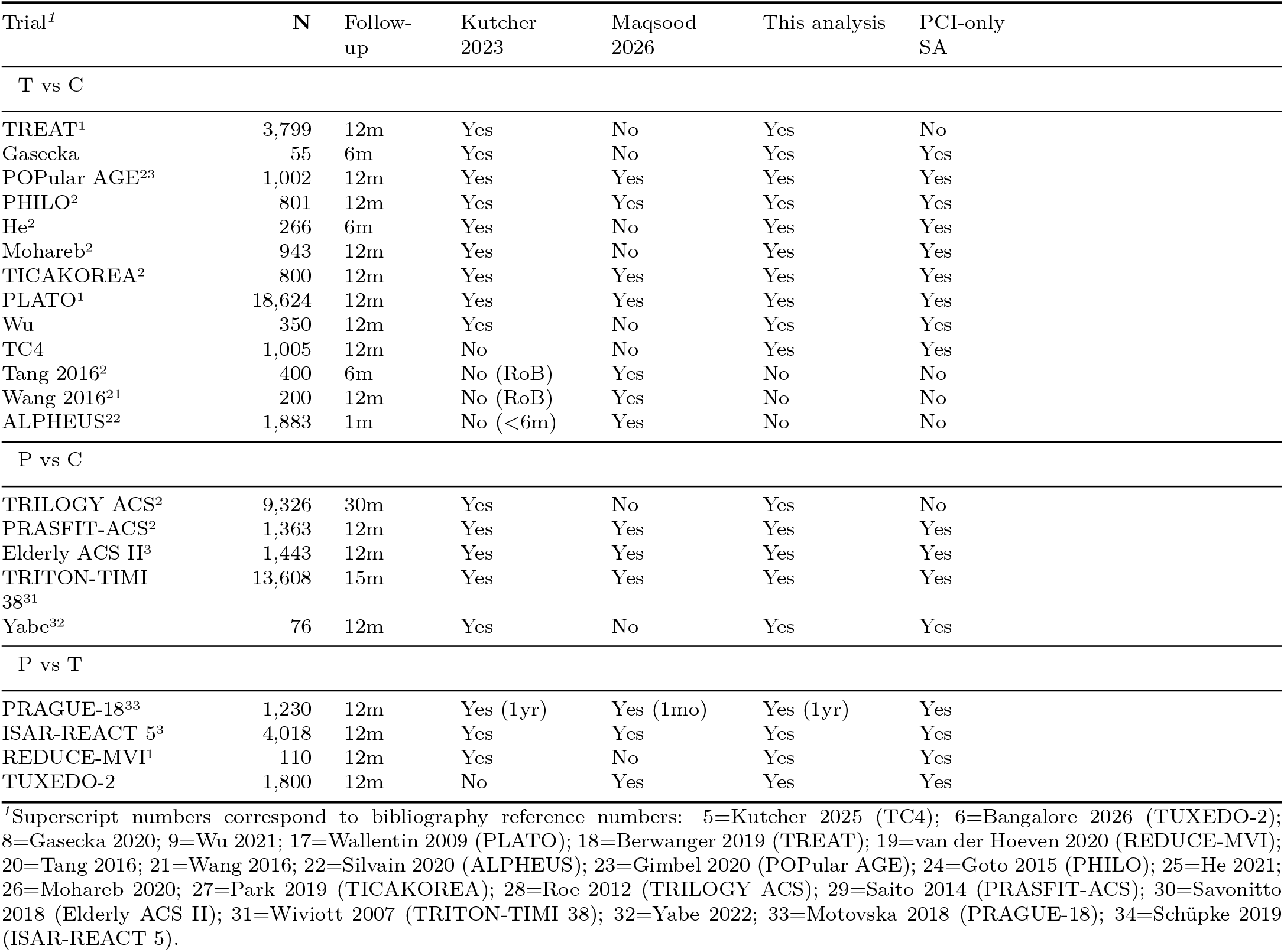
Trial reconciliation across Kutcher 2023, Maqsood 2026, and current analysis.

**Table 2:** Table 2. Trial-level MACE input data. Hard MACE = all-cause death + non-fatal MI + non-fatal stroke. Rate = events/n (%). For trials with no clopidogrel arm (P vs T), the reference arm is ticagrelor.

| Trial<sup>f</sup> | Follow-up | Reference arm | Events/n (%) | Comparator arm | Events/n (%) |
| --- | --- | --- | --- | --- | --- |
| P vs T |  |  |  |  |  |
| ISAR-REACT 5<sup>3</sup> | 12m | T (ref) | 208/2012 (10.3%) | P | 152/2006 (7.6%) |
| PRAGUE-18<sup>33,2</sup> | 12m | T (ref) | 56/634 (8.8%) | P | 44/596 (7.4%) |
| REDUCE-MVI<sup>1</sup> | 12m | T (ref) | 7/54 (13.0%) | P | 5/56 (8.9%) |
| TUXEDO-2 | 12m | T (ref) | 90/901 (10.0%) | P | 77/899 (8.6%)<sup>3</sup> |
| C vs P |  |  |  |  |  |
| Elderly ACS II<sup>3</sup> | 12m | C | 60/730 (8.2%) | P | 57/713 (8.0%) |
| PRASFIT-ACS<sup>2</sup> | 12m | C | 89/678 (13.1%) | P | 75/685 (10.9%) |
| TRILOGY ACS<sup>2 4</sup> | 30m | C | 854/4663 (18.3%) | P | 808/4663 (17.3%) |
| TRITON-TIMI 38<sup>31</sup> | 15m | C | 877/6795 (12.9%) | P | 724/6813 (10.6%) |
| Yabe<sup>32</sup> | 12m | C | 2/39 (5.1%) | P | 2/37 (5.4%) |
| C vs T |  |  |  |  |  |
| Gasecka | 6m | C | 0/28 (0.0%) | T | 0/27 (0.0%) |
| He<sup>2</sup> | 6m | C | 4/133 (3.0%) | T | 5/133 (3.8%) |
| Mohareb<sup>2</sup> | 12m | C | 9/472 (1.9%) | T | 4/471 (0.8%) |
| PHILO<sup>2</sup> | 12m | C | 31/400 (7.8%) | T | 43/401 (10.7%) |
| PLATO<sup>1</sup> | 12m | C | 1205/9291 (13.0%) | T | 1028/9333 (11.0%) |
| POPular AGE<sup>23</sup> | 12m | C | 79/500 (15.8%) | T | 81/502 (16.1%) |
| TC4 | 12m | C | 64/555 (11.5%) | T | 50/450 (11.1%) |
| TICAKOREA<sup>2</sup> | 12m | C | 31/400 (7.8%) | T | 42/400 (10.5%) |
| TREAT<sup>1</sup> | 12m | C | 164/1886 (8.7%) | T | 154/1913 (8.1%) |
| Wu | 12m | C | 0/174 (0.0%) | T | 0/176 (0.0%) |

<sup>f</sup>Superscript numbers correspond to bibliography reference numbers: 5=Kutcher 2025 (TC4); 6=Bangalore 2026 (TUXEDO-2); 8=Gasecka 2020; 9=Wu 2021; 17=Wallentin 2009 (PLATO); 18=Berwanger 2019 (TREAT); 19=van der Hoeven 2020 (REDUCE-MVI); 23=Gimbel 2020 (POPular AGE); 24=Goto 2015 (PHILO); 25=He 2021; 26=Mohareb 2020; 27=Park 2019 (TICAKOREA); 28=Roe 2012 (TRILOGY ACS); 29=Saito 2014 (PRASFIT-ACS); 30=Savonitto 2018 (Elderly ACS II); 31=Wiviott 2007 (TRITON-TIMI 38); 32=Yabe 2022; 33=Motovska 2018 (PRAGUE-18); 34=Schüpke 2019 (ISAR-REACT 5).
<sup>2</sup>PRAGUE-18: 1-year outcomes used (Motovska 2018, JACC 71:371), not 1-month (Circulation 2016). 39% crossover to clopidogrel by 1 year.
<sup>3</sup>TUXEDO-2 MACE: secondary endpoint 'death + MI + stroke' (Table 2, Bangalore 2026); primary endpoint also included major bleeding. Italicised to distinguish from trials where hard MACE was primary.
<sup>4</sup>TRILOGY ACS: 30-month follow-up; 0% PCI (medically managed UA/NSTEMI only). Excluded from PCI-only sensitivity analysis.

**Table 3:**
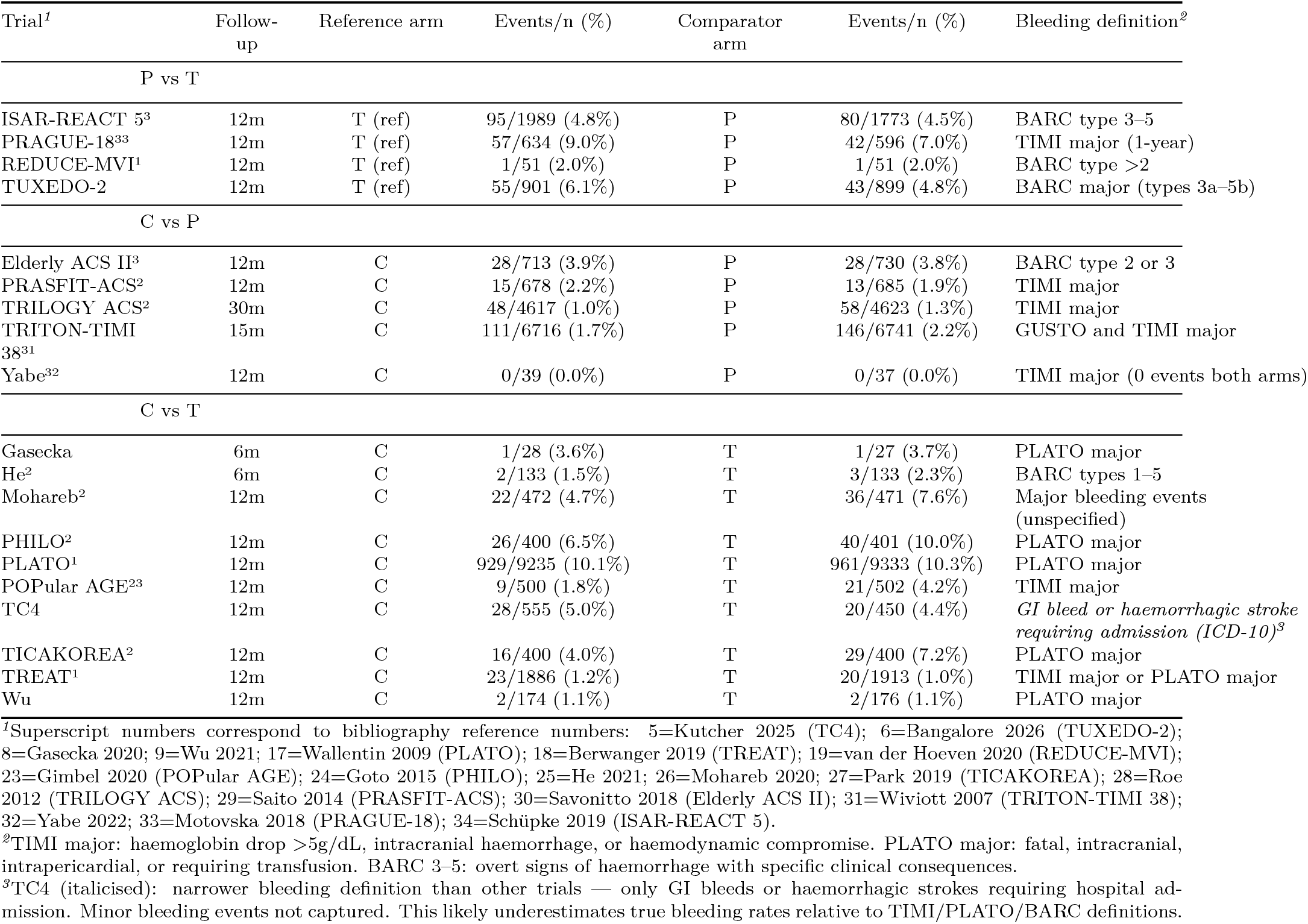
Table 3. Major bleeding input data. Bleeding definitions vary across trials; see footnotes. Rate shown as events/n (%).

**Figure 1:**
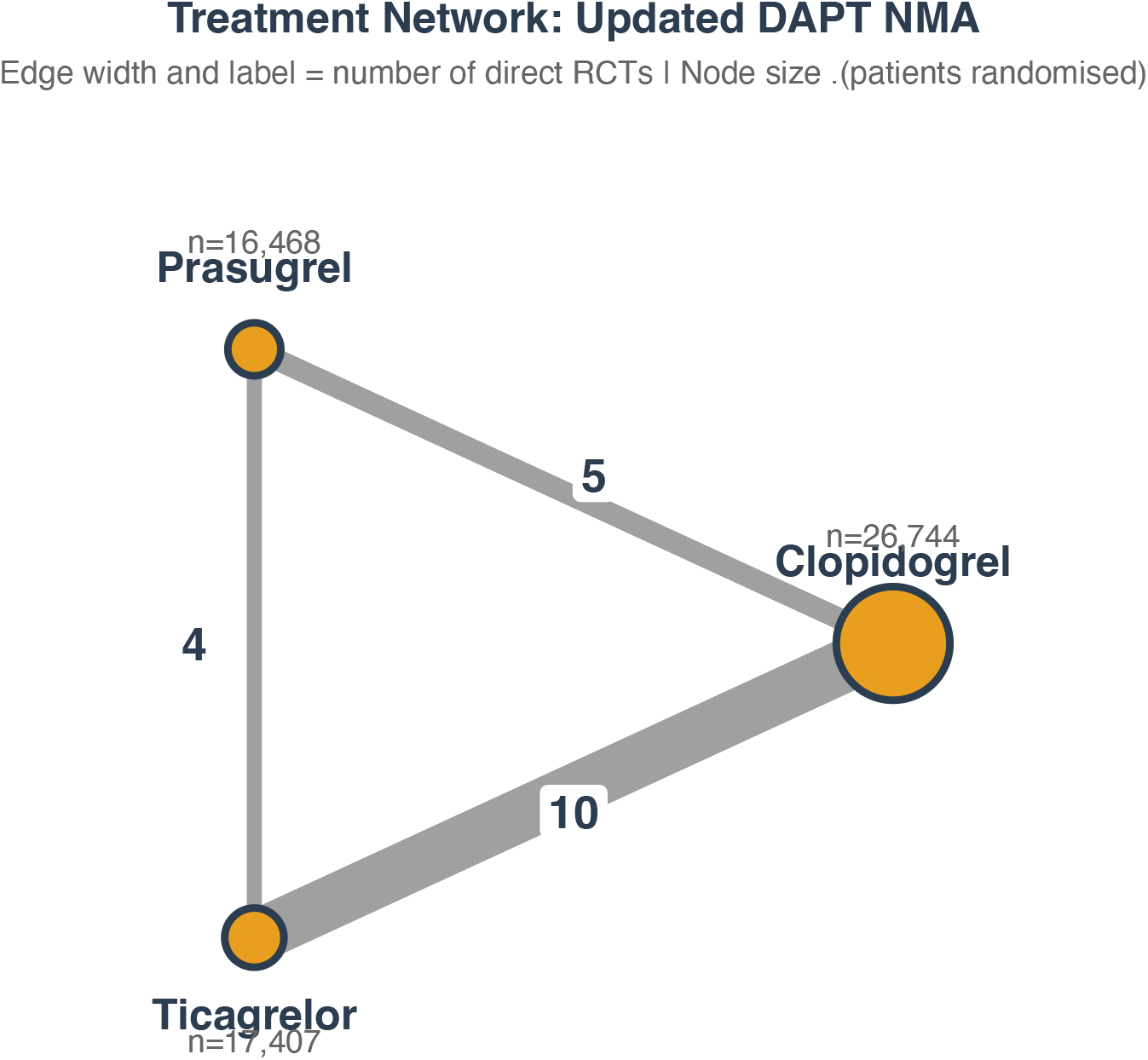
Network geometry for the updated Bayesian NMA (19 trials, n 60,619). Node size proportional to total patients randomised to that treatment across all trials. Edge width and label show number of direct RCTs per comparison. Nodes represent treatments; edge widths are proportional to the number of trials making each direct comparison; edge labels show trial counts. The triangle is fully connected — all three pairwise comparisons have direct RCT evidence — which minimises reliance on indirect inference. The T vs C edge carries the most trials (10) and the most patients (∼40,000), dominated by PLATO. The P vs T edge has the fewest direct trials (4) but includes the two largest head-to-head comparisons (ISAR-REACT 5, n = 4,018; TUXEDO-2, n = 1,800).

### 3.2 Prior Predictive Checks and Parameter Recovery

Given the importance of priors in a Bayesian analysis, we first performed prior predictive checks to assess model plausibility. This asks, before seeing the actual data, what event rates does the model consider plausible? The data contributes only the structural skeleton (study IDs, treatment levels, and follow-up times) needed to define the parameter space dimensions. Supplemental Figure 1 shows that the model reliably generates plausible prior event rates. A second preliminary step simulated data from known true parameters and confirmed that the model correctly recovered them, verifying the inferential machinery (data not shown).

### 3.3 Main Models

Primary random effects models were fitted for both MACE and bleeding outcomes and results are shown in Table 4. Overall this Bayesian network meta-analysis found no definitive evidence for the clinical superiority of any specific drug. Prasugrel did show a 70% probability of clinical benefit compared to clopidogrel (HR 0.87, 95% CrI 0.76– 1.03), a meaningful signal but with genuine uncertainty, as the CrI crosses 1.0. Prasugrel also shows some clinical benefit compared to ticagrelor (HR 0.89, 95% CrI 0.73–1.12) but this 95% CrI includes both clinically meaningful benefit and clinically meaningful harm and the associated 54% probability of a clinical benefit (HR < 0.9) is best described as marginal. Ticagrelor shows only very weak evidence for any superiority over clopidogrel (HR 0.98, 95%CrI 0.82–1.18) with 76% probability of falling in the ROPE, suggesting the most defensible conclusion for this comparison is one of clinical equivalence.

**Table 4:** Main results: Bayesian NMA^1^. 19 triais, n ≈ 60,619 | Primary model: binomial cloglog, log(time) offset, random effects

| Table 4. Main Results: Bayesian NMA<sup>1</sup> |  |  |  |  |
| --- | --- | --- | --- | --- |
| 19 trials, n ≈ 60,619 \| Primary model: binomial cloglog, log(time) offset, random effects |  |  |  |  |
| Comparison | HR (95% CrI)<sup>2</sup> | Pr(HR < 0.90) Benefit<sup>3</sup> | Pr(0.90–1.11) Equivalence | Pr(HR > 1.11) Harm |
| **MACE (cloglog RE)** |  |  |  |  |
| Ticagrelor vs Clopidogrel | 0.98 (0.82–1.18) | 16.8% | 75.5% | 7.6% |
| Prasugrel vs Clopidogrel | 0.87 (0.76–1.03) | 69.7% | 29.6% | 0.7% |
| Prasugrel vs Ticagrelor | 0.89 (0.73–1.12) | 54.0% | 43.0% | 3.0% |
| **Major Bleeding (cloglog RE)** |  |  |  |  |
| Ticagrelor vs Clopidogrel | 1.26 (1.01–1.63) | 0.3% | 13.4% | 86.3% |
| Prasugrel vs Clopidogrel | 1.11 (0.88–1.36) | 3.5% | 45.6% | 50.9% |
| Prasugrel vs Ticagrelor | 0.88 (0.67–1.14) | 57.8% | 39.0% | 3.2% |

<sup>1</sup> HR: hazard ratio; CrI: credible interval; ROPE: region of practical equivalence [0.90, 1.11]. Comparisons use full drug names: Ticagrelor (T), Clopidogrel (C), Prasugrel (P).
<sup>2</sup> HR: hazard ratio from cloglog model with log(time) offset. CrI: Bayesian credible interval. ROPE: region of practical equivalence [0.90, 1.11].
<sup>3</sup> Pr(HR < 0.90): probability of clinically meaningful benefit (>10% relative reduction). Pr(HR > 1.11): probability of clinically meaningful harm (>11% relative increase). Thresholds match Kutcher 2023 and TC4 (Kutcher 2025).

The clearest safety signal is increased bleeding with ticagrelor compared to clopidogrel (HR 1.26, 95%CrI 1.01–1.63), an 86% probability of meaningful harm. Prasugrel also exhibited increased bleeding compared to clopidogrel but decreased bleeding compared to ticagrelor. The efficacy and safety results are shown graphically in Figure 2.

**Figure 2:**
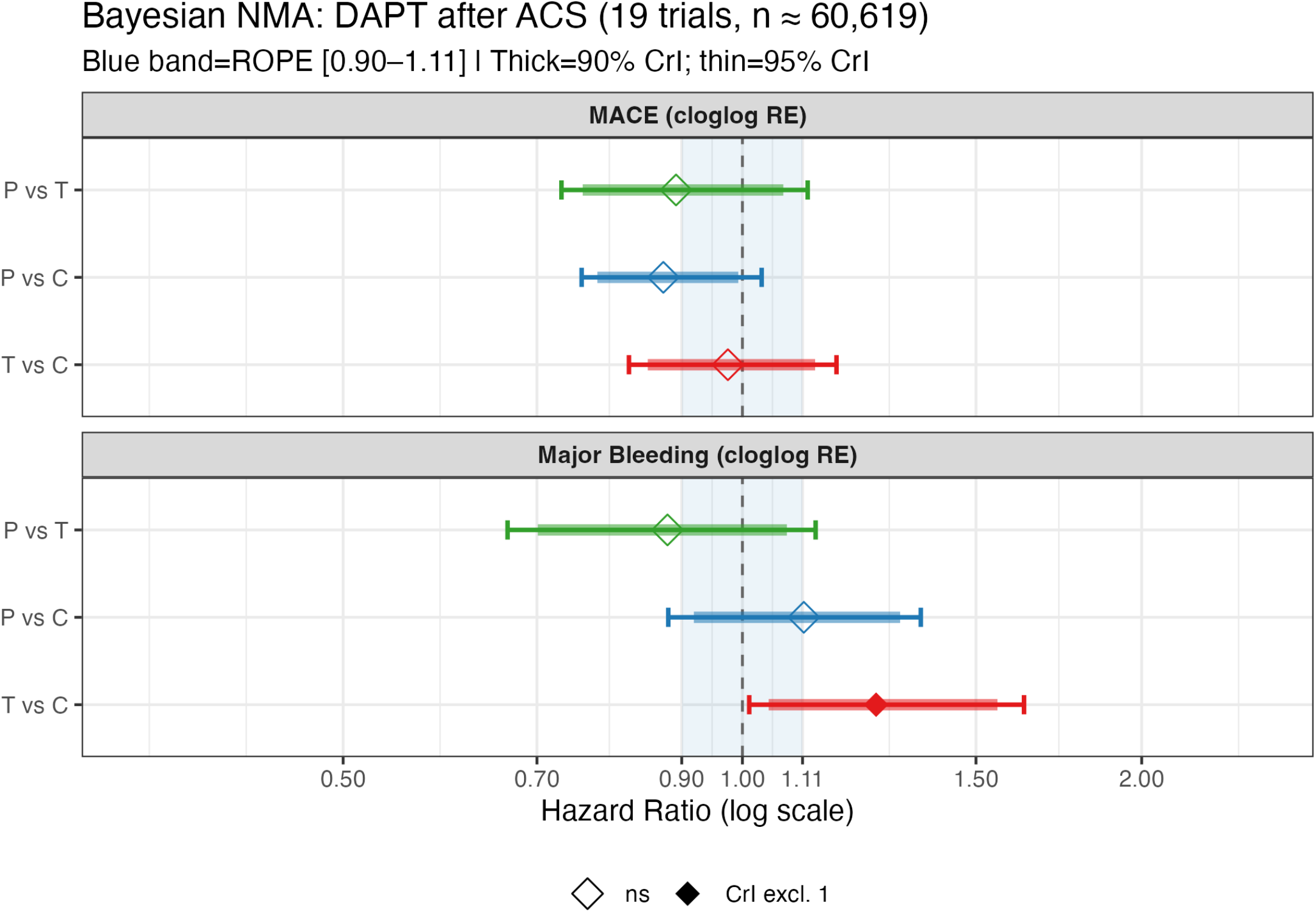
Figure 2. Bayesian network meta-analysis of P2Y12 inhibitor therapy after ACS (19 RCTs, n 60,619): posterior hazard ratio distributions for MACE (upper panel) and major bleeding (lower panel). Points represent the posterior median hazard ratio; thick and thin horizontal bars represent the 90% and 95% credible intervals, respectively. The shaded blue band denotes the region of practical equivalence (ROPE: HR 0.90–1.11), within which differences are considered clinically negligible. A filled diamond () indicates that the 95% CrI excludes 1.0. Hazard ratios below 1.0 favour the first-named treatment. C = clopidogrel; P = prasugrel; T = ticagrelor; HR = hazard ratio; CrI = credible interval; ROPE = region of practical equivalence.

The models showed excellent convergence with 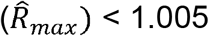 and no divergences. Sensitivity analyses showed the random effects model, which allows study-specific treatment effect deviations *u*_*ij*_, outperformed the fixed effect models that constrain treatment effects to be homogeneous across all trials (Leave-one-out cross-validation (LOO-CV), elpd_diff = −13.2, SE = 5.8, ratio = 2.3). Seven sensitivity analyses were performed as detailed in Supplemental Table 1 and Supplemental Figure 2. Results were robust to all sensitivity challenges. We also observed good consistency for the prasugrel versus ticagrelor efficacy comparison using node-splitting, comparing the direct trial estimates(6,33–35) (HR 0.79, 95%CrI 0.63 – 1.06) with the combined direct / indirect estimate derived from the Bayesian network meta-analysis (HR 0.89, 95%CrI 0.73 – 1.12) but with improved precision.

## 4. Discussion

After integrating the totality of evidence with a principled accounting of the associated uncertainties, our primary result is that prasugrel, with a 70% probability of clinically meaningful MACE benefit over clopidogrel and 54% over ticagrelor, represents the strongest efficacy signal in the network, though with genuine residual uncertainty. Its 51% probability of meaningful bleeding harm versus clopidogrel is not negligible and must be weighed against the efficacy gains. Prasugrel is contraindicated in those with prior stroke or TIA and should be used cautiously in patients aged ≥75 years or weighing <60 kg. Outside these restrictions in low bleeding risk post-PCI patients it likely provides the best net benefit.

Ticagrelor is the weakest performer. Its MACE benefit over clopidogrel is clinically negligible in the average patient (76% of posterior in ROPE), while its bleeding penalty is real and consistent (86% probability of meaningful harm). North American evidence (PLATO NA subgroup HR 1.25(17), TC4 RR 0.95(5)) consistently showed no efficacy advantage. Twice-daily dosing and common side-effects (dyspnea) are further disadvantages.

Clopidogrel performs at least as well as ticagrelor on MACE and substantially better on bleeding. The clinical efficacy gap between clopidogrel and prasugrel is also fairly uncertain. As a once-daily generic with a favourable safety profile and good adherence data, clopidogrel appears undervalued by current guidelines — particularly for older patients, those at elevated bleeding risk, and North American patients where ticagrelor’s global trial result(17) has not been replicated in regional or dedicated RCTs and prasugrel has not been adequately studied.

Why does our MACE estimate for prasugrel versus ticagrelor (HR 0.89, 95% CrI 0.73– 1.12, 54% probability of benefit) differ markedly from the JAMA network meta-analysis(3) estimate (OR 0.83 (95%CI 0.70–0.98, p = 0.03))? Five factors may contribute to explaining this discrepancy.

First, the JAMA comparison rests on direct evidence from three trials(6,34,36) and the indirect evidence flowing through the clopidogrel node. A fourth direct prasugrel versus ticagrelor trial (19) has been added to this analysis. However this trial is small (n=110) and contributes little weight, so this difference alone does not explain the discrepancy. More importantly the new analysis treats one of the direct trials, PRAGUE-18 differently. We used 1-year outcomes (33) rather than the 1-month outcomes(36) as in the JAMA NMA(3). At 1 month, the prasugrel advantage was higher but by 1 year, with 39% crossover to clopidogrel, the apparent prasugrel benefit was attenuated. Using 1-year data is methodologically correct for a 12-month treatment comparison but substantially reduces the PRAGUE-18 contribution to prasugrel’s advantage.

Second, the indirect estimate of the prasugrel versus ticagrelor hazard ratio is derived mathematically as the difference between the prasugrel versus clopidogrel log-hazard ratio and the ticagrelor versus clopidogrel log-hazard ratio. When the ticagrelor versus clopidogrel estimate moves toward equivalence (log-HR toward zero), the indirect prasugrel versus ticagrelor estimate narrows accordingly. Since TC4(5) showed near-equivalence of ticagrelor and clopidogrel, and the PLATO North American subgroup(17) showed no ticagrelor benefit, the pooled estimate is pulled toward HR = 1.0. This narrows the gap between prasugrel and ticagrelor in the indirect path, reducing the apparent prasugrel advantage in the combined network estimate. The Bayesian random effects model propagates this heterogeneity explicitly with less weight accorded to the indirect contribution to the prasugrel versus ticagrelor estimate. The JAMA paper(3) does not account for this heterogeneity propagation in the same way.

Third, with only four direct prasugrel versus ticagrelor trials, the between-study SD *τ* is estimated with much uncertainty, reflecting genuine incertitude about how much the true effect varies across populations. This uncertainty has two consequences. First, the credible interval for the population-average treatment effect *β*_*j*_ widens relative to what a fixed effects model produces, because uncertainty in τ propagates into uncertainty about the treatment effect *β*_*j*_. Second, the predictive interval for what the true effect would be in a new trial population is wider still, incorporating both the uncertainty in *β*_*j*_ and the expected between-trial variability. In contrast, a fixed effects model achieves a narrower interval by assuming *τ* = 0 (no heterogeneity), which is an implausible assumption across four different populations (Japan/Korea, Germany, the Netherlands, India). The random effects model’s wider interval is the honest representation of how much four heterogeneous trials can inform.

Fourth the JAMA paper(3) used SUCRA (surface under the cumulative ranking curve) to rank treatments This is computed from point estimates and ignores associated uncertainties. SUCRA can therefore assign a high rank to a treatment even when confidence intervals substantially overlap. The 54% posterior probability of clinical benefit in the current analysis is a more direct and interpretable statement of the actual evidence — it explicitly quantifies that there is nearly a 50% chance the true prasugrel versus ticagrelor advantage does not reach the clinically meaningful threshold.

Finally, the JAMA paper(3) reported odds ratios without adjusting for follow-up time, despite individual trials varying between one(36) and 12 months(6,34) follow-up. When follow-up is uniform, the OR and HR are numerically similar, but the current model’s time offset ensures that even small differences in actual follow-up within the 12-month trials (due to censoring and loss to follow-up) are correctly handled. More importantly, the OR systematically overestimates the HR when event rates are above 10% — ISAR-REACT 5’s ticagrelor arm had 10.3% MACE — so the frequentist OR slightly overstates the magnitude of any prasugrel advantage

This Bayesian approach has several strengths. First, it avoids a dichotomisation of uncertainty. For example, the JAMA NMA(3) frames the prasugrel versus clopidogrel as “significant” (p < 0.05) and ticagrelor versus clopidogrel as “non-significant” (p > 0.05), implying a qualitative difference where the data are actually compatible with a quantitative one. The Bayesian posterior distributions show that 70% vs 17% probability of clinically meaningful MACE benefit is a difference of degree, not a binary distinction. Second, it facilitates the construction of ROPE, making clinical equivalence an explicit conclusion. For ticagrelor versus clopidogrel MACE, 76% of the posterior lies within HR [0.90, 1.11]. This is not a failure to detect a difference — it is a positive finding that the drugs are clinically interchangeable for this endpoint in the average trial population. While this interpretation is self evident within the Bayesian paradigm, it is obscured in the frequentist analysis. Third, the Bayesian approach propagates between study heterogeneity into estimates in a principled manner and rejects a default of homogenous effects as scientifically untenable. The Bayesian model standardizes all trials to a common hazard rate, making the pooling statistically coherent. Finally, seven sensitivity analyses have confirmed robustness across alternative model specifications, link functions, trial inclusions, and prior choices.

After integrating the totality of evidence across 19 RCTs and approximately 60,000 patients, the honest summary which become explicit under a Bayesian workflow is one of modest, uncertain, and context-dependent treatment differences. One large direct trial(34) provided clear evidence for prasugrel superiority over ticagrelor, but three other direct trials were inconclusive(6,19,33). When pooled with appropriate random effects uncertainty and the network indirect evidence — which is shaped by the heterogeneous ticagrelor versus clopidogrel results — the Bayesian estimate is that prasugrel is probably better than ticagrelor for MACE, but the probability of a clinically meaningful difference (HR < 0.90) is only 54%, not strong enough to support a definitive hierarchy. Clinical decision-making should therefore be individualised: prasugrel is a reasonable preference in high-thrombotic-risk patients without contraindications, but a universal categorical recommendation as previously suggested(3) is not warranted. Perhaps the most important clinical conclusion of this analysis is that clopidogrel — unfairly demoted by guidelines anchored to a single global trial result that has not been replicated in North American or dedicated regional RCTs — deserves reconsideration as a first-line option, particularly for older patients and those at elevated bleeding risk.

## Data Availability

All data produced are available online at https://github.com/brophyj/Bayesian-NMA

## 7 Supplemental material

### 7.1 Figures

**Figure 3: Supplemental Figure 1.**
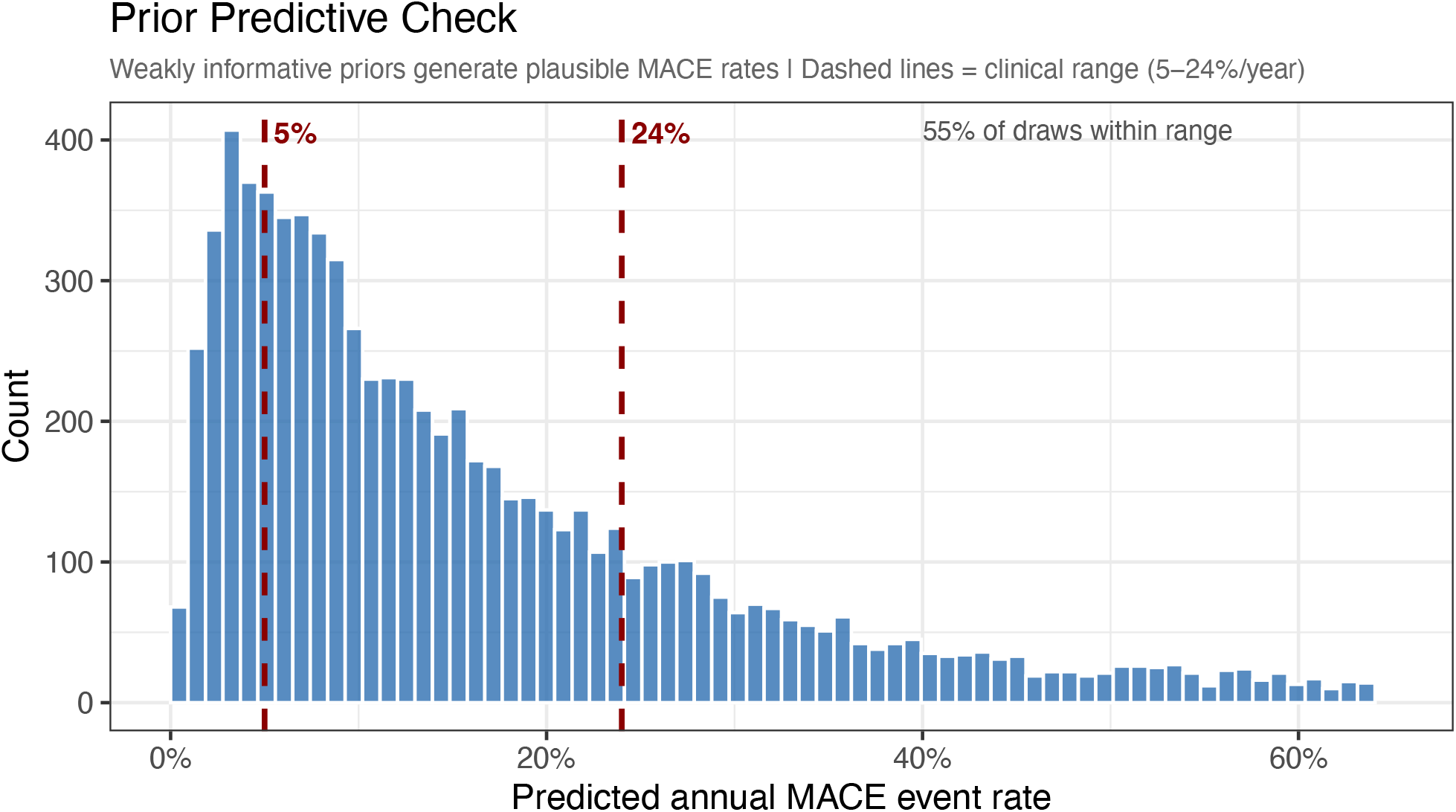
Prior predictive check: predicted annual MACE rates under the weakly informative priors. Dashed lines mark the clinically plausible range (5–24%/year).

**Supplemental Figure 2:**
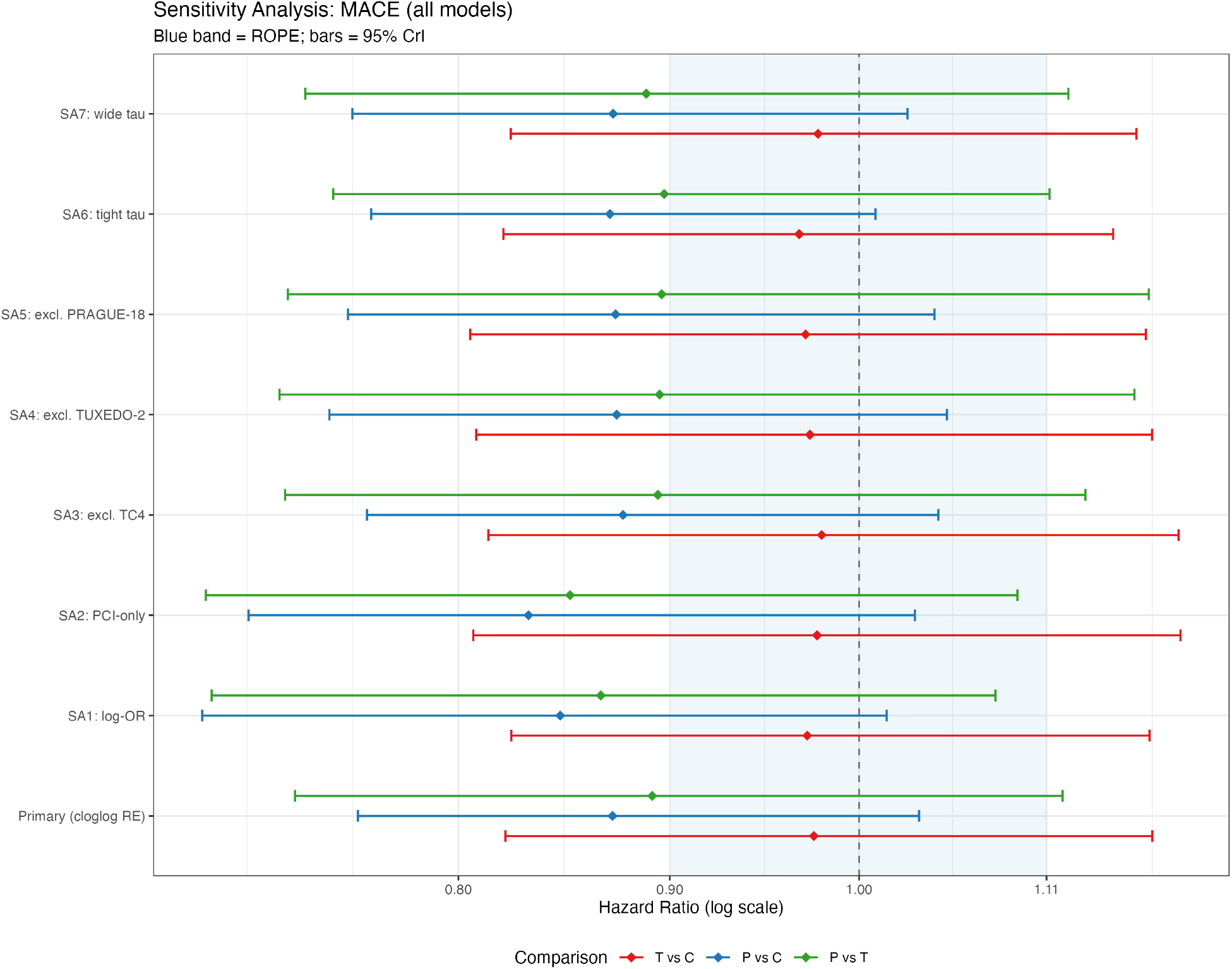
Forest plot of the sensitivity analyses. *C = clopidogrel; P = prasugrel; T = ticagrelor. SA = sensitivity analysis. SA1: log-OR model; SA2: PCI-only trials; SA3: excluding TC4; SA4: excluding TUXEDO-2; SA5: excluding PRAGUE-18; SA6: tighter heterogeneity prior; SA7: wider heterogeneity prior. HR = hazard ratio; CrI = credible interval; ROPE = region of practical equivalence [0.90, 1.11].*

### 7.2 Tables

| Sensitivity Analyses: MACE<sup>†</sup> |  |  |  |  |
| --- | --- | --- | --- | --- |
| All models: binomial cloglog RE unless noted |  |  |  |  |
| Comparison | HR (95% CrI) | Pr(benefit) | Pr(equivalence) | Pr(harm) |
| **Primary (cloglog RE)** |  |  |  |  |
| T vs C | 0.98 (0.82–1.18) | 16.8% | 75.5% | 7.6% |
| P vs C | 0.87 (0.76–1.03) | 69.7% | 29.6% | 0.7% |
| P vs T | 0.89 (0.73–1.12) | 54.0% | 43.0% | 3.0% |
| **SA1: log-OR** |  |  |  |  |
| T vs C | 0.97 (0.82–1.18) | 17.9% | 74.8% | 7.3% |
| P vs C | 0.85 (0.69–1.02) | 80.7% | 18.7% | 0.6% |
| P vs T | 0.87 (0.70–1.08) | 64.9% | 33.6% | 1.5% |
| **SA2: PCI-only** |  |  |  |  |
| T vs C | 0.98 (0.81–1.20) | 18.2% | 72.0% | 9.8% |
| P vs C | 0.83 (0.71–1.03) | 83.1% | 16.1% | 0.8% |
| P vs T | 0.85 (0.69–1.09) | 70.3% | 27.7% | 2.0% |
| **SA3: excl. TC4** |  |  |  |  |
| T vs C | 0.98 (0.81–1.19) | 17.6% | 73.0% | 9.4% |
| P vs C | 0.88 (0.76–1.05) | 66.2% | 33.0% | 0.8% |
| P vs T | 0.89 (0.73–1.13) | 52.6% | 44.0% | 3.3% |
| **SA4: excl. TUXEDO-2** |  |  |  |  |
| T vs C | 0.97 (0.81–1.18) | 19.3% | 72.7% | 8.0% |
| P vs C | 0.87 (0.74–1.05) | 67.3% | 31.5% | 1.3% |
| P vs T | 0.89 (0.72–1.17) | 52.2% | 43.4% | 4.4% |
| **SA5: excl. PRAGUE-18** |  |  |  |  |
| T vs C | 0.97 (0.81–1.17) | 19.6% | 72.9% | 7.5% |
| P vs C | 0.87 (0.75–1.04) | 68.9% | 30.1% | 1.0% |
| P vs T | 0.90 (0.73–1.18) | 52.0% | 43.2% | 4.8% |
| **SA6: tight tau** |  |  |  |  |
| T vs C | 0.97 (0.82–1.15) | 18.5% | 75.5% | 6.0% |
| P vs C | 0.87 (0.76–1.01) | 71.6% | 28.1% | 0.3% |
| P vs T | 0.90 (0.75–1.11) | 51.2% | 46.2% | 2.6% |
| **SA7: wide tau** |  |  |  |  |
| T vs C | 0.98 (0.82–1.17) | 16.3% | 76.1% | 7.5% |
| P vs C | 0.87 (0.75–1.03) | 68.9% | 30.4% | 0.7% |
| P vs T | 0.89 (0.73–1.12) | 55.3% | 41.6% | 3.1% |

<sup>†</sup> SA1: logistic (logOR) model, no time offset. SA2: PCI-only trials (excludes TREAT and TRILOGY ACS). SA3: excludes TC4 (cluster-RCT design). SA4: excludes TUXEDO-2 (selected DM+MVD population). SA5: excludes PRAGUE-18 (39% crossover to clopidogrel at 1 year). SA6: tighter between-study SD prior [half-Normal(0, 0.25)]. SA7: wider between-study SD prior [half-Normal(0, 1.0)].

## Footnotes

1 **Derivation of the half-Normal mean.** If *Z* ∼ Normal (0, 1), the standard normal is symmetric around zero so half its mass lies below zero and half above. Taking the absolute value |*Z*| reflects the negative half onto the positive axis — this is the folding operation that produces the half-normal. The factor of 2 below captures this reflection explicitly, doubling the integral over the positive half to account for the mass we folded across: 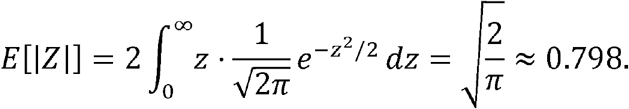 Now, a Norm (0, *σ*) random variable *X* is simply a rescaled standard normal X = *σZ*, so |*X*| = *σ*|*Z*|. Taking expectations, 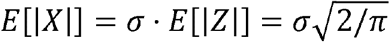. With *σ* = 0.5, this gives *E*[*τ*] = 0.5 × 0.798 ≈ 0.40 — the 0.5 enters because *τ* is drawn from Normal (0,0.5), not Normal(0,1), and the half-normal mean scales linearly with *σ*.

## Notes

Conflicts of interest: The authors have no conflicts of interest to report.

### Competing Interest Statement

The authors have declared no competing interest.

### Author Declarations

Uses only previously published data

